# Regression Analysis of COVID-19 Spread in India and its Different States

**DOI:** 10.1101/2020.05.29.20117069

**Authors:** Poonam Chauhan, Ashok Kumar, Pooja Jamdagni

## Abstract

Linear and polynomial regression model has been used to investigate the COVID-19 outbreak in India and its different states using time series epidemiological data up to 26^th^ May 2020. The data driven analysis shows that the case fatality rate (CFR) for India (3.14% with 95% confidence interval of 3.12% to 3.16%) is half of the global fatality rate, while higher than the CFR of the immediate neighbors *i.e*. Bangladesh, Pakistan and Sri Lanka. Among Indian states, CFR of West Bengal (8.70%, CI: 8.21–9.18%) and Gujrat (6.05%, CI: 4.90–7.19%) is estimated to be higher than national rate, whereas CFR of Bihar, Odisha and Tamil Nadu is less than 1%. The polynomial regression model for India and its different states is trained with data from 21^st^ March 2020 to 19^th^ May 2020 (60 days). The performance of the model is estimated using test data of 7 days from 20^th^ May 2020 to 26^th^ May 2020 by calculating RMSE and % error. The model is then used to predict number of patients in India and its different states up to 16^th^ June 2020 (21 days). Based on the polynomial regression analysis, Maharashtra, Gujrat, Delhi and Tamil Nadu are continue to remain most affected states in India.

## Introduction

Since the outbreak of COVID-19 from the Wuhan city of China, the novel corona virus has gripped more than 200 countries around the globe and the number of patients and causality have been risen up to 52,04,508 and 3,37,687, respectively, as on 24^th^ May 2020 [1]. On looking at the rate of spread and threat for human life, the World Health Organization (WHO) declared COVID-19 a pandemic on 11 March 2020 [2]. The first case of COVID-19 in India was found on 30 January 2020 in the state of Kerala in a student who had returned to home from Wuhan University, China and soon after the epidemic spread in the other part of country mostly due to the imported cases from outside the country. The outbreak of COVID-19 (SARS-CoV-2) in India and its different states has caused illness in 1,31,867 patients and 3867 deaths as on 25^th^ May 2020 [3].

The symptoms COVID-19 include high fever, dry cough, body pain and respiratory distress. Its incubation period varies from 2 to 14 days and the most important mode of its transmission is respiratory droplets and contact transmission [4]. Some patients are also found to be asymptotic i.e. they do not show any symptoms [5]. Such cases are silent spreaders of virus and are most dangerous and difficult to trace out. In such situation a random testing may play important role to contain the spread of the virus at community level. So far in India, the community spread is at the cluster level in the cities like Mumbai, Chennai and Ahmedabad with number of patients as large as 30,500, 10,500 and 10,300, respectively [6].

In order to contain the spread of virus and to prevent its human-to-human transmission, Indian government had announced a nation-wide lockdown of 21 days from the midnight of 24th March 2020 and subsequent series of lockdowns of 19 days and 15 days. The 4^th^ phase of Lockdown 4.0, has started from 18^th^ of May 2020. Since these measures have brought huge pressure on economy, therefore, to revive the economy, Prime minister Modi has announced a package of Rs. 20 lakh crores (20 trillion) on 12^th^ May 2020. In the absence of a COVID-19 vaccine at the moment the only accepted way to attenuate the growth is to practice good hand hygiene, using masks compulsorily, restricting public gatherings, increasing quarantine facility, increasing dedicated COVID-19 hospitals, increasing sample testing and follow social distancing.

Although, several studies in the context of India have been reported recently by many researchers [7–15] to understand and analyze the dynamics of COVID-19 spread, but there are very limited studies on state wise analysis [16–18] of the outbreak. Looking at the diversity in population, population density and geographical conditions, the study of India as a whole may not provide actual status of the epidemic, therefore, each states of India which has huge populations as compared to the other part of world, need to analyze separately for the spread of coronavirus.

Statistical models are important tools to analyze the real-time data analysis of infectious disease. In this paper, we have utilized the linear and polynomial regression model to analyze the epidemic data of India and its different states. The short-term forecast of the expected patients in next three weeks are also estimated using polynomial regression. It is important to mention that the prediction made in this study is as good as the quality of data available and deviation from the trends in coming days may change the predictions as well.

## Method

The epidemiological data of COVID-19 cases was collected from official website of Ministry of Health and Family Welfare, Government of India (https://www.mohfw.gov.in). We apply simple linear regression model to estimate case fatality rate (CFR) and recovery rate (RR) using data up to 26^th^ May 2020 for Indian and its different states. We use cumulative number of confirmed cases as predictor variable and cumulative deaths or recovered cases as outcome variable. The day of reporting first death case or recovered case in each state was used as starting point in the linear regression model, doing so we discard the influence on CFR and RR due to initial stage outbreak with no deaths or recovery. The coefficient of determination (R^2^) is evaluated to determine the good fit of the model. The slope of the fitted line has been used to estimate the CFR and RR. The 95% confidence interval (CI) is calculated from the standard error of the slope. Also, the polynomial regression model is used to forecast the number of expected patients in next three weeks up to 16^th^ June 2020. The least square polynomial has been determined for each state and the results are validated with statistical error analysis. Initial 90% data (60 days from March 21,2020 to May 19, 2020) was used for training the model and remaining 10% data (7 days from May 20, 2020 to May 26, 2020) for test purpose for validation of the model by calculating RMSE and % error. The trained and tested model is then used to predict number of patients from 27^th^ May 2020 to 16^th^ June 2020 in India and its different states.

## Results and Discussion

The simple linear regression analysis of number of deaths as a function of number of confirmed cases for India is shown in Figure 1(a). The coefficient of determination (R^2^) is calculated to be 0.997 which implies strong linear correlation between confirmed and death cases reported up to 26^th^ May 2020 from the first death case. The case fatality rate (CFR) is estimated to be 3.14% with 95% confidence interval 3.12% to 3.16%. CFR of India is half of global fatality rate (6.38%), as on 26^th^ May 2020 [19]. Note that the CFR (as on 26^th^ May 2020) of most COVID-19-affected countries such as USA, Brazil, Spain, UK, Italy, France, Turkey and Iran is 5.95%, 6.24%, 11.40%, 14.18%, 14.26%, 19.57%, 4.62%, 5.47%, respectively, which is higher than India, whereas the CFR of Russia and Germany is lower (1.03% and 2.77%, respectively) than India [19]. The CFR of immediate neighbors Bangladesh, Pakistan and Sri Lanka is 1.43%, 2.07% and 0.78%, respectively, which is better than India [19].

**Figure 1:**
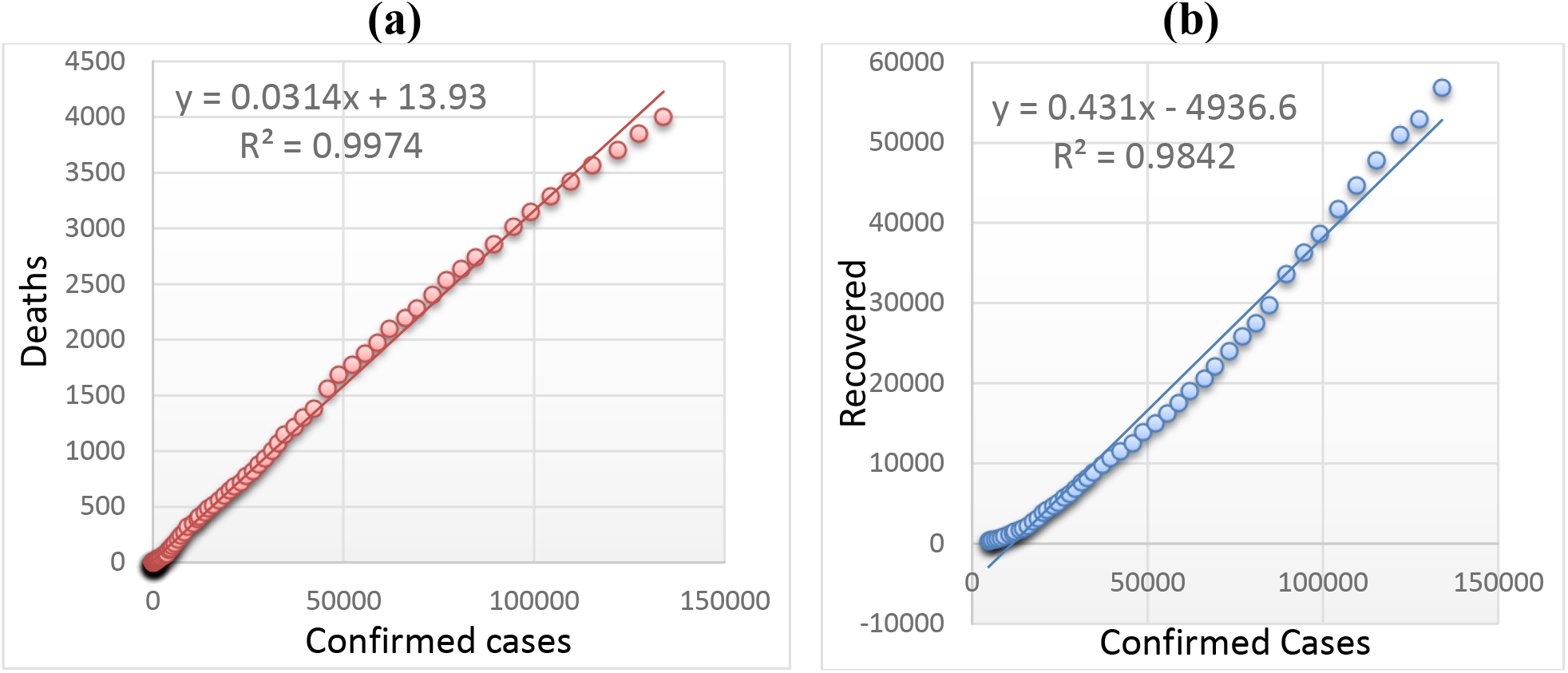
Linear regression model analysis of total number of cases with (a) total number of deaths (b) total number of recovered cases, in India (data taken up to 26^th^ May 2020).

The simple linear regression analysis of number of recovered cases as a function of number of confirmed cases for India is shown in Figure 1(b). The calculated R^2^ of 0.984 implies linear correlation between confirmed and recovered cases (as on 26^th^ May 2020). The recovery rate (RR) is estimated to be 43.1% with 95% confidence interval 41.4% to 44.7%, as compared to global 43.3% (as on 26^th^ May 2020). The RR (as on 26^th^ May 2020) of top virus hit countries such as USA, Brazil, Italy, France and Russia is 28.1%, 39.8%, 39.7%, 36.4% and 39.8% respectively, which is lower than India, whereas Spain (69.1%), Turkey (77.2%), Iran (78.5%) and Germany (89.5%) has RR much higher than world as well as India [20]. It is also important to mention that the outbreak of pandemic in Italy, USA, Spain, Iran and Germany was earlier than India. The RR of immediate neighboring countries of India *i.e*. Bangladesh, Pakistan and Sri Lanka are 20.9%, 33% and 48.9%, respectively, as on 26^th^ May 2020 [20].

The number of deaths/recovered cases as a function of total confirmed cases for different states of India are shown in Figure 2 and Supplementary Information. R^2^ for confirmed cases versus death (recovered) cases for different states varies from 0.897 to 0.994 (0.894 to 0.988). The CFR of West Bengal (8.70%, CI: 8.21–9.18%), Gujrat (6.05%, CI: 4.90–7.19%), Madhya Pradesh (4.82%, CI: 4.59–5.05%) and Maharashtra (3.40%, CI: 3.37–3.50%) is estimated to be higher than national rate *i.e*. 3.14 %, whereas CFR of Bihar, Odisha and Tamil Nadu is less than 1%. The RR of Telangana and Maharashtra is estimated to be highest and lowest with 88.6% (CI: 79.3–97.8%) and 27.7% (CI: 26.3–29.1%), respectively. CFR, RR and statistical parameters for different states are listed in Table 1. Considering the ongoing national and state wise variation in the number of cases, the CFR and RR is subject to change in the later stage.

**Figure 2:**
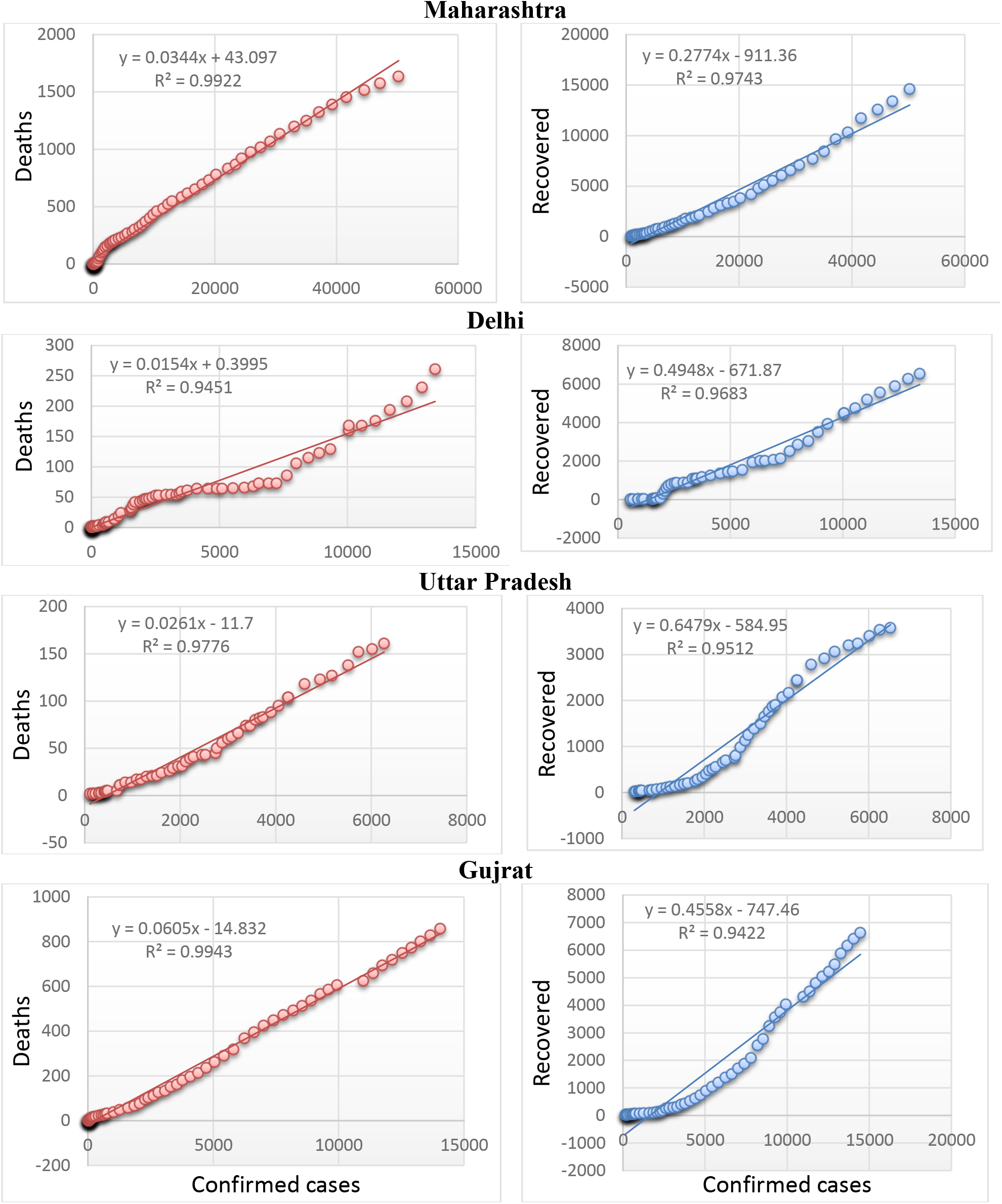
Linear regression model analysis of total number of cases with total number of deaths and recovered cases, in few representative states in India (data taken up to 26^th^ May 2020).

**Table 1:** Case fatality rate (CFR) and recovery rate (RR) estimation for India and its different states (for data up to 26^th^ May 2020) using linear regression model. Statistical parameters (95% confidence interval and R^2^) are also given.

| States | Case Fatality Rate |  |  | Recovery Rate |  |  |
| --- | --- | --- | --- | --- | --- | --- |
| | CFR (%) | 95% CI | $R^2$ | RR (%) | 95% CI | $R^2$ |
| **India** | **3.14** | **3.12-3.16** | **0.997** | **43.1** | **41.4-44.7** | **0.984** |
| Jammu & Kashmir | 1.14 | 1.09-1.18 | 0.965 | 58.1 | 56.01-60.1 | 0.988 |
| Haryana | 1.44 | 1.37-1.5 | 0.973 | 66.6 | 61.2-71.9 | 0.943 |
| Uttar Pradesh | 2.60 | 2.48-2.71 | 0.977 | 64.9 | 59.8-69.9 | 0.951 |
| Delhi | 1.50 | 1.44-1.63 | 0.945 | 49.4 | 46.3-52.4 | 0.968 |
| Rajasthan | 2.81 | 2.64-2.97 | 0.963 | 66.1 | 62.1-70.0 | 0.968 |
| Bihar | 0.50 | 0.47-0.52 | 0.959 | 29.9 | 23.6-36.1 | 0.913 |
| Madhya Pradesh | 4.82 | 4.59-5.05 | 0.973 | 63.1 | 58.4-67.7 | 0.959 |
| Andhra Pradesh | 2.07 | 0.91-2.18 | 0.964 | 73.2 | 66.4-79.9 | 0.929 |
| Odisha | 0.48 | 0.45-0.50 | 0.956 | 33.9 | 31.1-36.6 | 0.944 |
| Telangana | 2.54 | 2.4-2.6 | 0.967 | 88.6 | 79.3-97.8 | 0.911 |
| West Bengal | 8.70 | 8.21-9.18 | 0.964 | 36.8 | 34.4-39.1 | 0.960 |
| Maharashtra | 3.40 | 3.37-3.50 | 0.992 | 27.7 | 26.3-29.1 | 0.974 |
| Karnataka | 2.60 | 2.43-2.88 | 0.897 | 38.3 | 33.9-42.7 | 0.894 |
| Tamil Nadu | 0.64 | 0.61-0.66 | 0.983 | 43.0 | 38.3-47.6 | 0.908 |
| Gujrat | 6.05 | 4.90-7.19 | 0.994 | 45.0 | 42.6-47.3 | 0.942 |

Next, we consider the analysis of polynomial regression model for India and its different states. The first case of COVID-19 in India was reported on 30^th^ Jan 2020, thereafter, in the month of February only three cases were reported. The substantial rise in the cases in India were reported after 20^th^ march 2020. The number of confirmed patients as a function of number of days since 21^st^ March 2020 is shown in Figure 3(a). The data from 21^st^ March 2020 to 19^th^ May 2020 (60 days) is used to train the regression model. The best fit for epidemiological data of India is estimated for least square third-degree polynomial with R^2^ = 0.999 as shown in Figure 3(a). The model is subsequently tested with data from 20^th^ May 2020 to 26^th^ May 2020 (7days) and validated with root mean square error (RMSE). The predicted patients for next three weeks from 27^th^ May 2020 are plotted in Figure 3(b). The % error between actual and predicted data is estimated to be 2%. The number of patients by 16^th^ June 2020 is estimated to be 306666.

**Figure 3:**
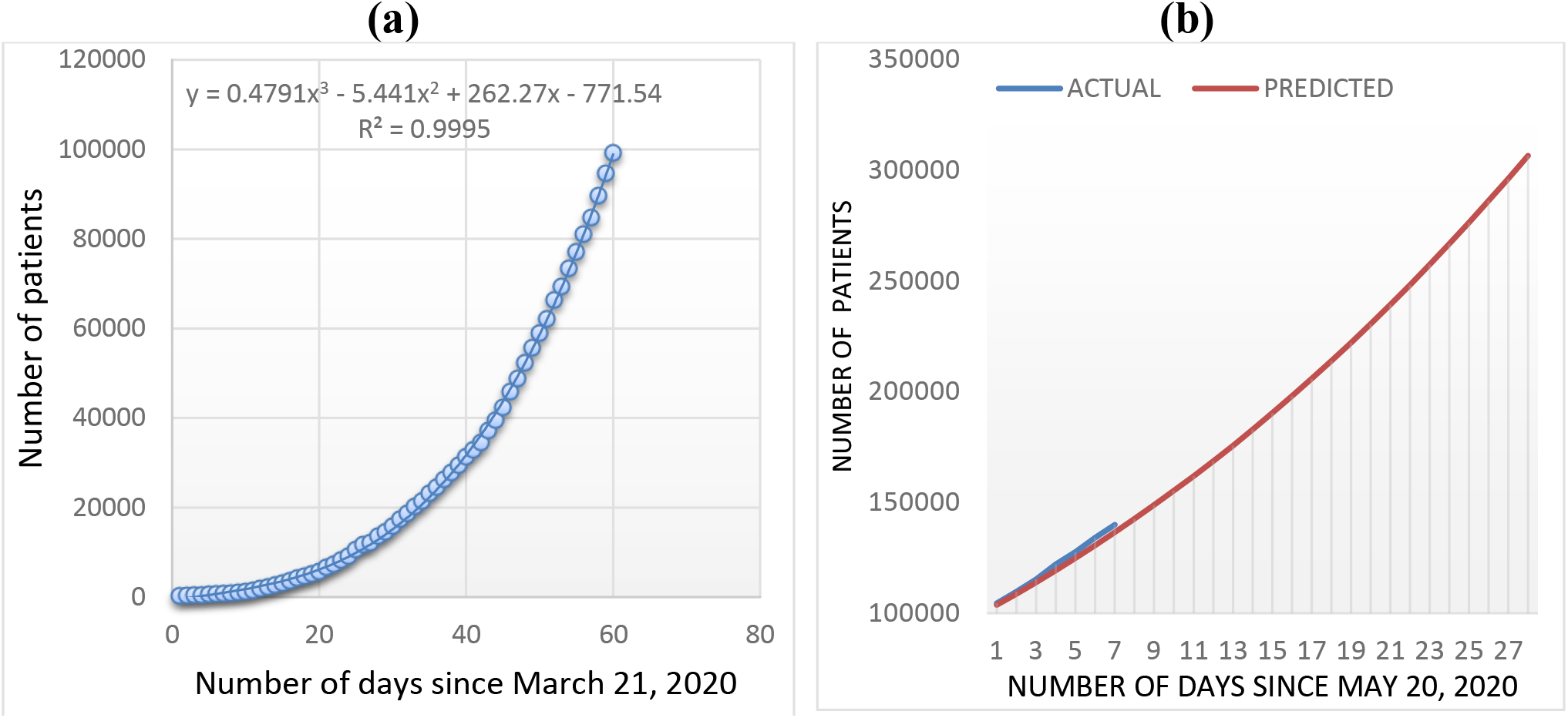
Polynomial regression model analysis for India. (a) Total number of cases as a function of training data set since March 21, 2020. (b) Total number of cases with test data from May 20, 2020 to May 26, 2020. Predicted patients from May 27, 2020 to June 16, 2020 are also plotted.

The number of confirmed patients as a function of number of days since 21^st^ March 2020 for different states are shown in Figure 4 and Supplementary Information. The value of R^2^ varies from 0.936 to 0.999. The degree of least square polynomial for different states varies from 2 to 3. The estimated number of patients in Maharashtra, Delhi, Gujrat and West Bengal are 119091, 32473, 29305 and 7174 with R^2^ (% error) of 0.999 (4%), 0.998 (2%), 0.998 (3%) and 0.995 (3%), respectively. The expected number of patients by 16^th^ June 2020, degree of least square polynomial, R^2^, RMSE and % error for different states are listed in Table 2. Based on the polynomial regression analysis, Maharashtra, Gujrat, Delhi and Tamil Nadu are continue to remain most affected states in India till 16^th^ June 2020.

**Figure 4:**
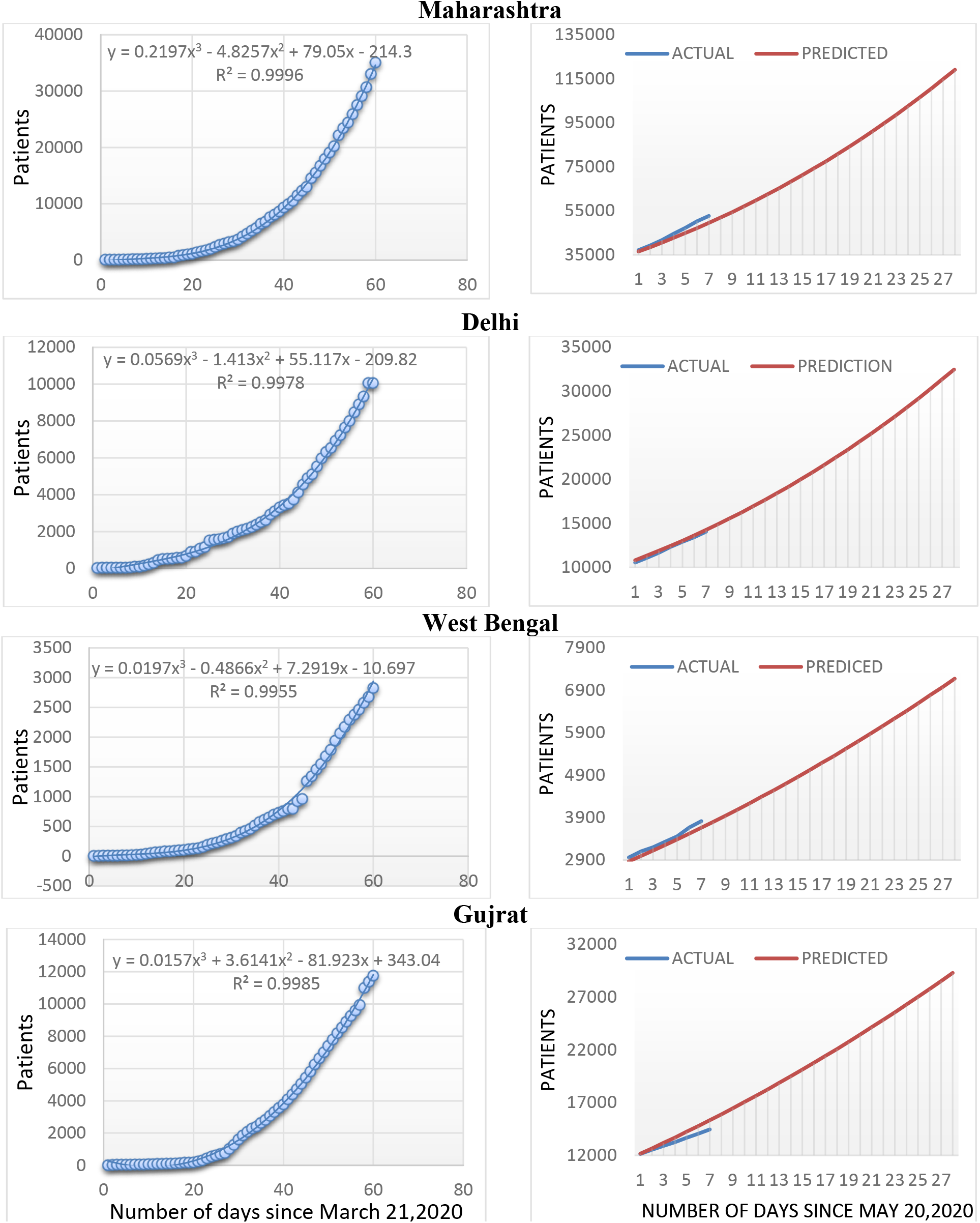
Polynomial regression model analysis for few representative states: (a) Total number of cases as a function of training data set since March 21, 2020. (b) Total number of cases as a function of test data from May 08, 2020 to May 19, 2020. Predicted patients from May 20, 2020 to June 02, 2020 are also plotted.

**Table 2:** Results of polynomial regression model with predicted number of infected peoples in India and its various states. Statistical parameters (R^2^, root mean square error and % error) are also given.

| State | Degree of polynomial | $R^2$ | % Error | RMSE | Predicted number of patients till 16<sup>th</sup> June 2020 |
| --- | --- | --- | --- | --- | --- |
| **India** | **3** | **0.999** | **2** | **2199** | **306666** |
| Andhra Pradesh | 3 | 0.997 | 2 | 70 | 3373 |
| Bihar | 3 | 0.986 | 24 | 545 | 4822 |
| Delhi | 3 | 0.998 | 2 | 198 | 32473 |
| Gujrat | 2 | 0.998 | 3 | 435 | 29305 |
| Haryana | 2 | 0.977 | 2 | 20 | 2085 |
| Jammu & Kashmir | 3 | 0.995 | 10 | 171 | 2520 |
| Karnataka | 3 | 0.994 | 23 | 464 | 2980 |
| Kerala | 3 | 0.991 | 14 | 128 | 1127 |
| Madhya Pradesh | 2 | 0.991 | 6 | 385 | 10865 |
| Maharashtra | 3 | 0.999 | 4 | 1856 | 119091 |
| Odisha | 3 | 0.970 | 6 | 68 | 4408 |
| Punjab | 2 | 0.936 | 35 | 705 | 6088 |
| Rajasthan | 2 | 0.997 | 8 | 524 | 11354 |
| Tamil Nadu | 3 | 0.988 | 9 | 1405 | 55233 |
| Telangana | 2 | 0.973 | 10 | 183 | 2075 |
| Uttar Pradesh | 2 | 0.996 | 8 | 483 | 9978 |
| West Bengal | 2 | 0.995 | 3 | 105 | 7174 |

## Conclusions

In summary, regression analysis was carried out to investigate the COVID-19 outbreak in India and its different states. Linear regression model is used to estimate case fatality rate (CFR) and recovery rate (RR) in India and its different states using time-series epidemiological data up to 26^th^ May 2020. CFR for India is estimated to be 3.14% with 95% confidence interval of 3.12% to 3.16%. The recovery rate (RR) of India is estimated to be 43.1% with 95% confidence interval of 41.4% to 44.7%, as compared to global 43.3% (as on 25^th^ May 2020). Among Indian states, CFR of West Bengal, Gujrat, Madhya Pradesh and Maharashtra is estimated to be higher than national rate, whereas CFR of Bihar, Odisha and Tamil Nadu is less than 1%. The RR of Telangana and Maharashtra is estimated to be highest and lowest, respectively. The polynomial regression model is used to predict the number of patients in next three week. The model is trained with data from 21^st^ March 2020 to 19^th^ May 2020 (60 days) and validated with test data of 7 days from 20^th^ May 2020 to 26^th^ May 2020 and calculating RMSE and % error. The predicted patients in India up to 16^th^ June 2020 can reach as large as 3 lakhs. Based on the polynomial regression analysis, Maharashtra, Gujrat, Delhi and Tamil Nadu are continue to remain most affected states in India till 16^th^ June 2020.

## Data Availability

Supplementary Information available for this paper.

## Acknowledgements

We acknowledge Prof. P. K. Ahluwalia, Himachal Pradesh University, Shimla for valuable discussion during the course of study.

## References

1. Situation report-125: Available at https://www.who.int/emergencies/diseases/novelcoronavirus2019/situationreports/?gclid=EAIaIQobChMIwsvj1IXN6QIVSqaWCh2THwWdE AAYASACEgLRAvD_BwE

2. Elmousalami, H.H. and A.E. Hassanien, Day level forecasting for Coronavirus Disease (COVID-19) spread: analysis, modeling and recommendations. arXiv preprint arXiv:2003.07778, 2020.

3. https://www.mohfw.gov.in/dashboard/index.php. Accessed on May 25, 2020

4. T. Singhal, A review of coronavirus disease-2019 (COVID-19). The Indian Journal of Pediatrics (2020) 87, 281–286

5. He, G., et al., The clinical feature of silent infections of novel coronavirus infection (COVID‐19) in Wenzhou. Journal of Medical Virology, 2020, **DOI:** 10.1002/jmv.25861

6. https://www.covid19india.org/

7. P. Kumar et. al., Forecasting COVID-19 impact in India using pandemic waves Nonlinear Growth Models, medRxiv 2020: https://doi.org/10.1101/2020.03.30.20047803

8. M. K. Prakash et. al., A minimal and adaptive prediction strategy for critical resource planning in a pandemic, medRxiv 2020: https://doi.org/10.1101/2020.04.08.20057414

9. J. S.Virk et. al., Recent update on COVID-19 in India: Is locking down the country enough? medRxiv 2020: https://doi.org/10.1101/2020.04.06.20053124

10. R. Singh et. al., Age-structured impact of social distancing on the COVID-19 epidemic in India, arXiv preprint 2020: arXiv:2003.12055v1 [q-bio.PE]

11. S. Mandal et. al., Prudent public health intervention strategies to control the coronavirus disease 2019 transmission in India: A mathematical model-based approach, Indian J. Med. Res (2020): DOI:10.4103/ijmr.IJMR_504_20

12. T. Sardar et. al., Assessment of 21 Days Lockdown Effect in Some States and Overall India: A Predictive Mathematical Study on COVID-19Outbreak, arXiv preprint 2020: arXiv:2004.03487v1 [q-bio.PE]

13. S. Ghosal et. al. Linear Regression Analysis to predict the number of deaths in India due to SARS-CoV-2 at 6 weeks from day 0 (100 cases – March 14^th^ 2020), Diabetes & Metabolic Syndrome: Clinical Research & Reviews (2020) 14 311–315

14. S. Yadav and P. K. Yadav, Basic Reproduction Rate and Case Fatality Rate of COVID-19: Application of Meta-analysis, medRxiv 2020: https://doi.org/10.1101/2020.05.13.20100750

15. R. Gupta, et. al., SEIR and Regression Model based COVID-19 outbreak predictions in India, medRxiv 2020: https://doi.org/10.1101/2020.04.01.20049825

16. P. Ghosh et. al., COVID-19 in India: State-wise Analysis and Prediction, medRxiv 2020: https://doi.org/10.1101/2020.04.24.20077792

17. N. Poonia and S. Azad, Short-term forecasts of COVID-19 spread across Indian states until 1 May 2020, https://arxiv.org/abs/2004.13538

18. S. Mondal et. al., Possibilities of exponential or Sigmoid growth of Covid19 data in different states of India, medRxiv 2020: https://doi.org/10.1101/2020.04.10.20060442

19. https://github.com/owid/covid-19-data/tree/master/public/data

20. https://www.worldometers.info/coronavirus/#countries

